# Stool processing methods for Xpert Ultra testing in childhood tuberculosis: A prospective, multi-country accuracy study

**DOI:** 10.1101/2024.12.17.24317956

**Authors:** Devan Jaganath, Pamela Nabeta, Mark P. Nicol, Robert Castro, Peter Wambi, Heather J. Zar, Lesley Workman, Rakesh Lodha, Urvashi B. Singh, Ashish Bavdekar, Sonali Sanghavi, André Trollip, Aurélien Mace, Maryline Bonnet, Manon Lounnas, Petra de Haas, Edine Tiemersma, David Alland, Padmapriya Banada, Adithya Cattamanchi, Morten Ruhwald, Eric Wobudeya, Claudia M. Denkinger, the Stool H2H Study Team

**Affiliations:** Division of Pediatric Infectious Diseases, University of California, San Francisco, San Francisco, USA; Center for Tuberculosis, University of California, San Francisco, San Francisco, USA; FIND, Geneva, Switzerland; Marshall Centre, School of Biomedical Sciences, University of Western Australia, Perth, Australia; Division of Pulmonary and Critical Care Medicine, University of California, San Francisco, San Francisco, USA; Walimu, Kampala, Uganda; Department of Paedatrics and Child Health, University of Cape Town, Cape Town, South Africa; Department of Pediatrics, All India Institute of Medical Sciences, New Delhi, India; Department of Microbiology, All India Institute of Medical Sciences, New Delhi, India; Department of Pediatrics, KEM Hospital, Pune, India; Department of Microbiology, KEM Hospital, Pune, India; TransVIHMH, University of Montpellier, IRD, Inserm, Montpellier, France; University of Montpellier, IRD, CNRS, MIVEGEC, Montpellier, France; KNCV Tuberculosis Foundation, The Hague, The Netherlands; Public Health Research Institute, Department of Medicine, Rutgers-New Jersey Medical School, Newark, USA; Division of Pulmonary Diseases and Critical Care Medicine, University of California Irvine, Orange, USA; Department of Infectious Disease and Tropical Medicine, University Hospital Heidelberg, German Center of Infection Research, partner site Heidelberg, Germany

**Author notes:** <u>Corresponding Author</u> Devan Jaganath, MD MPH Department of Pediatrics University of California, San Francisco 550 16th Street, 4th Floor San Francisco, CA 94158 USA. Contributed equally as first authors (Devan Jaganath, Pamela Nabeta). Contributed equally as senior authors (Eric Wobudeya, Claudia M. Denkinger).

**Keywords:** child, tuberculosis, diagnostics, stool, Xpert Ultra, centrifuge-free

## Abstract

**Background:** Centrifuge-free processing methods support stool Xpert Ultra testing for childhood tuberculosis (TB), but there are limited data on their accuracy, acceptability and usability.

**Methods:** We conducted a prospective evaluation of stool Xpert Ultra in India, South Africa, and Uganda with three methods: Stool Processing Kit (SPK), Simple One-Step (SOS), and Optimized Sucrose Flotation (OSF). Children <15 years old with presumptive TB had respiratory specimen testing with Xpert Ultra and culture. Stool was tested using Xpert Ultra after processing with each method. We compared the accuracy of each method to a microbiological reference standard (MRS) and a composite reference standard (CRS). We surveyed the laboratory staff to assess acceptability and usability of the methods.

**Results:** We included 607 children, of whom the median age was 3.5 years (IQR 1.3-7), 48% were female, and 15.5% were HIV positive. Against the MRS, the sensitivities of SPK, SOS and OSF were 36.9% (95% CI 28.6-45.8), 38.6% (95% CI 17.2-51), and 31.3% (95% CI 20.2-44.1), respectively. The specificities of SPK, SOS and OSF were 98.2% (95% CI 96.4-99.3), 97.3% (95% CI 93.7-99.1) and 97.1% (95% CI 93.3-99), respectively. Laboratory staff reported that the methods were acceptable and usable, but SOS was most feasible to implement in a peripheral facility. Sensitivity increased among children who were culture-positive (55-77.3%) and was low (13-16.7%) against the CRS.

**Conclusions:** Stool processing methods for Xpert Ultra were acceptable, usable, and performed similarly, with highest sensitivity among children with culture-positive TB.

**KEY POINTS:** In a multi-country diagnostic accuracy study for childhood pulmonary tuberculosis, three stool processing methods for Xpert Ultra were acceptable, usable and performed similarly. Sensitivity was lower than that of sputum Xpert Ultra, but improved in children with culture-positive disease.

## INTRODUCTION

The implementation of low-complexity, automated molecular assays for *Mycobacterium tuberculosis (Mtb)* has improved access to tuberculosis (TB) testing worldwide [1]. However, children are often unable to expectorate sputum, and access to equipment and trained staff to collect induced sputum or gastric aspirates is usually limited to higher-level facilities [2].

Multiple studies have shown that *Mtb* can be detected in the stool of children with pulmonary TB [3–6], in particular with Xpert MTB/RIF and Xpert MTB/RIF Ultra (Xpert Ultra, Cepheid, Sunnyvale). To support stool Xpert Ultra testing, centrifuge-free stool processing methods have been developed, including the Stool Processing Kit (SPK, FIND, Geneva) [7], Simple One-Step method (SOS, KNCV TB Foundation, Hague, Netherlands) [8], and Optimized Sucrose Flotation method (OSF, TB SPEED Consortium) [9]. However, limited comparative data exist on their accuracy, acceptability and usability [10].

Our overall objective was therefore to evaluate and compare the diagnostic accuracy of stool processing methods for Xpert Ultra testing through a prospective, multi-country study. We also assessed their acceptability and usability as reported by laboratory staff.

## METHODS

### Setting and Participants

We consecutively enrolled children under 15 years old from June 2019 to March 2021 in India, South Africa, and Uganda. In India, children were recruited from two tertiary care centers: the All India Institute of Medical Sciences (AIIMS), New Delhi, and KEM Hospital, Pune, with referrals from surrounding outpatient clinics. In South Africa, children were recruited from two tertiary hospitals, Red Cross Children’s Hospital in Cape Town and Dora Nginza Hospital in Gqeberha. In Kampala, Uganda, children were referred from Mulago National Referral Hospital inpatient and outpatient facilities, and hospitals and clinics from the surrounding area. Enrollment was paused for periods during the COVID-19 pandemic, and each site followed local regulations of when they could resume. Children were eligible if they had microbiologically confirmed TB, or at least one symptom of pulmonary TB: unexplained cough for ≥2 weeks, unexplained fever for ≥1 week, unexplained failure to thrive or weight loss, or chest X-ray suggestive of TB. Children already on anti-TB treatment for >72 hours were excluded. Caregivers completed a written informed consent, and children aged ≥8 years in Uganda, and ≥7 years in South Africa and India gave assent according to local ethical guidelines. Laboratory staff aged ≥18 years who processed the stool samples at each site completed an anonymous survey to assess acceptability and usability.

The study received ethical approval from the Mulago Hospital Research Ethics Committee, the Human Research Ethics Committee of the University of Cape Town, the AIIMS Ethics Committee and the KEM Hospital Research Centre Ethics Committee, and the University of California San Francisco Institutional Review Board.

### Reference Procedures

Trained staff performed a standard TB evaluation on all children, including a clinical questionnaire, physical exam, and chest radiography. Two sputum specimens were collected, including expectorated or induced sputum, gastric aspirate or nasopharyngeal aspirate, and tested using Xpert Ultra and liquid or solid mycobacterial culture according to standard operating procedures. Sputum Xpert Ultra trace results were defined as positive [11].

### Stool Collection

Study staff asked participants to collect one stool sample in a sterile cup at enrollment, either directly or transferred from a diaper using a spoon. If the child was unable to produce a sample, the caregiver was provided with a cup and was asked to return the sample as soon as obtained within 3 days.

Stool samples were homogenized when possible and processed fresh, or stored at 2-8°C and tested within 72 hours. Based on available supplies, protocols and training, SPK was introduced first in June 2019, followed by SOS and then OSF in December 2019. When all three methods were tested, we randomized the order of testing and where each sample was collected in the cup each week.

### Stool processing and index testing

Stool samples were processed using the following methods before Xpert Ultra testing (**Supplemental Figure 1**):

1. <u>SPK</u>. The kit (42T for FIND) included a squeeze bottle with mixing beads, a stool processing buffer (SPB) and a filter cap. Five milliliters (mL) of the SPB and 5 mL of Xpert Ultra Sample Reagent (SR) were pipetted into the squeeze bottle. One gram of stool was added to the squeeze bottle, shaken vigorously, and incubated at room temperature for 30 minutes. The mixture was then squeezed through the filter cap into the Xpert Ultra cartridge.
2. <u>SOS:</u> 0.8-1 gram of solid stool was added to an 8 mL SR bottle. If the stool was soft or liquid, then respectively 2 or 4 mL of SR were replaced with the same volume of stool. Shaking and incubation for 10 minutes were performed twice, and then two mL of supernatant were pipetted into the Xpert Ultra cartridge.
3. <u>OSF:</u> 0.5 gram of stool was added to 10 mL of 50% Sheather’s solution, and emulsified using wooden sticks and manually shaking. After 30 minutes of incubation to allow sedimentation, 0.5 mL of supernatant were mixed with 1.8 mL of SR and incubated for 15 minutes before being added to the Xpert Ultra cartridge.

Of note, the three methods were at different stages of development. SOS was design-locked, while SPK was a prototype design, and OSF had not yet been developed into a kit.

Laboratory staff were blinded to the sputum Xpert Ultra result and clinical treatment decision. Within three months of the start of enrollment, the staff were asked to complete a voluntary, anonymous acceptability and usability questionnaire on each method, using a Likert scale ranging from Totally or Partially Disagree to Partially or Totally Agree.

### TB Classification

Participants were classified as Confirmed, Unconfirmed or Unlikely TB based on NIH consensus definitions [12]. Confirmed TB was defined as having sputum positive for *Mtb* by Xpert Ultra or culture. Children with Unconfirmed TB did not have microbiological confirmation, but had signs, symptoms and/or radiographic findings consistent with TB and had clinical response to anti-TB treatment. Children with Unlikely TB had symptom resolution without anti-TB treatment. A case was unclassifiable if there was insufficient information or follow up to determine TB status. Stool Xpert Ultra results were not used in the TB classification.

### Statistical Analysis

Our primary analysis assessed the sensitivity and specificity of stool Xpert Ultra using each method, against a microbiological reference standard (MRS) that classified TB in children with Confirmed TB, and otherwise as not having TB (Unconfirmed or Unlikely TB). Children who could not provide a sputum or stool sample were excluded. Participants with non-determinate stool Xpert Ultra results (invalid, error or no result) were excluded; however, we conducted a sensitivity analysis using an intention-to-diagnose approach that considered their results as either negative or positive. For the head-to-head comparison, we included children who had valid results for all three methods, and used McNemar’s test with 95% CIs to compare differences in sensitivity and specificity. Statistical significance was defined if 95% CIs of the difference did not cross zero.

Secondarily, we assessed accuracy using three additional reference standards: 1) a composite reference standard (CRS) that included Unconfirmed TB in the definition of TB; 2) sputum Xpert Ultra result alone; and 3) sputum culture results alone. In addition, we determined the incremental accuracy of using both stool and sputum Xpert Ultra compared to each test alone against sputum culture.

For the acceptability and usability assessment, we summarized the frequency and proportion of the responses by processing method. Analyses were completed using Stata version 16.1 (StataCorp LLC, College Station, Texas). Findings have been presented in accordance with the Standards for Reporting Diagnostic Accuracy (STARD) guidelines [13].

## RESULTS

### Participant Characteristics

We enrolled 655 children from 1,182 screened across the three countries (**Figure 1**). After excluding 29 children who did not provide a stool sample and 19 children whose TB status was unclassifiable, the final sample size was 607. Most children were from Uganda (n=371, 61.1%) and under 5 years old (n=367, 60.5%, **Table 1**). Of the 574 children, 89 (15.5%) were living with HIV and 308 (50.7%) were underweight. A total of 147 children (24.2%) had Confirmed TB, with the majority (136/147, 92.5%) being sputum Xpert Ultra positive.

**Figure 1.**
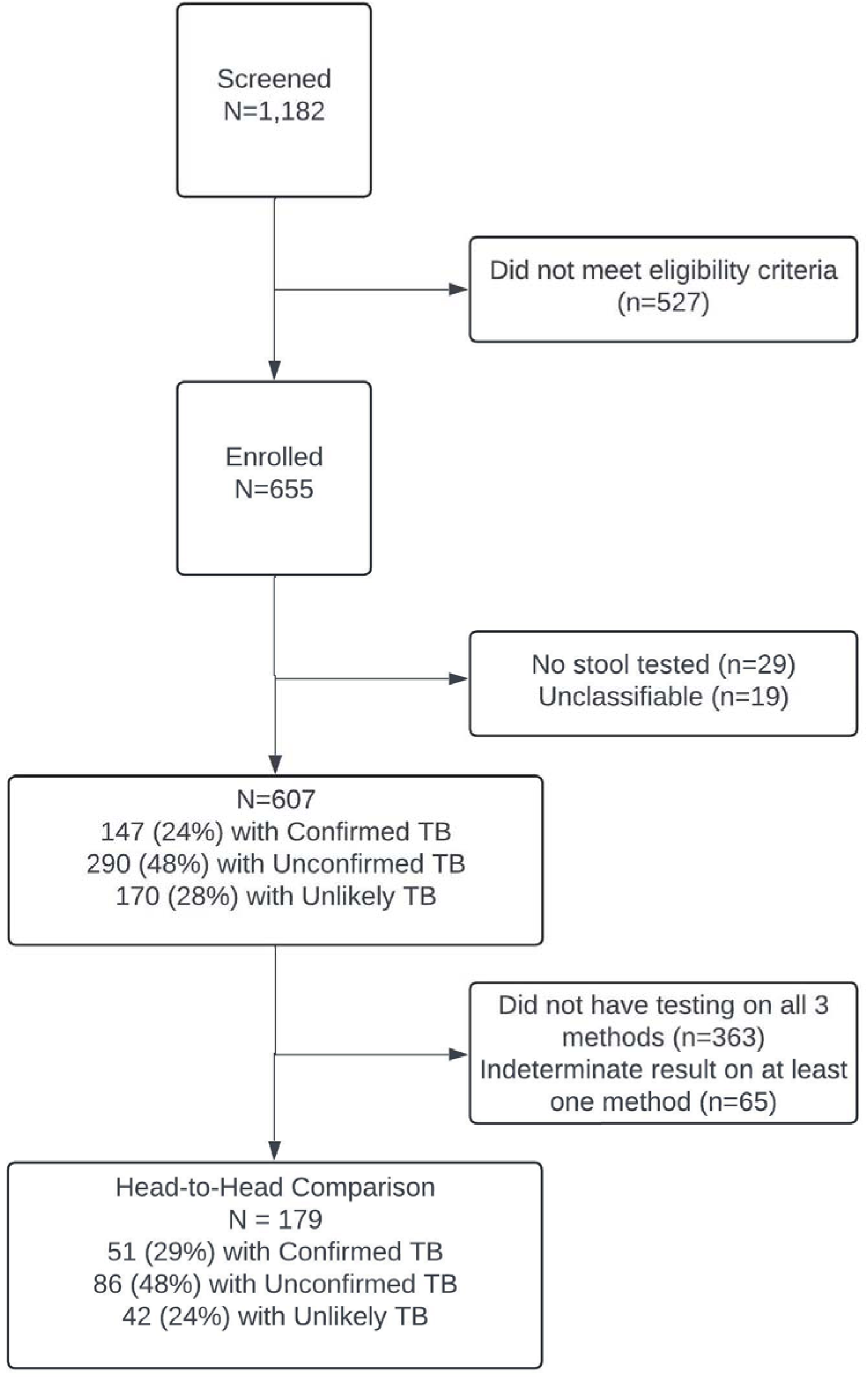
Flow of Participants.

**Table 1.** Sample Characteristics.

| **Characteristic<sup>a</sup>** | **Median (IQR) or n (%)** |
| --- | --- |
| Age | 3.5 (1.3-7) |
| <5 years old | 367 (60.5) |
| 5-9 years | 164 (27.0) |
| 10-14 years | 76 (12.5) |
| Female | 292 (48.1) |
| HIV positive (n=574) | 89 (15.5) |
| Median CD4 (n=72) | 715 (261-1228) |
| Underweight | 308 (50.7) |
| History of TB (n=604) | 19 (3.2) |
| History of known TB contact (n=602) | 314 (52.2) |
| Country |  |
| India | 58 (9.6) |
| South Africa | 178 (29.3) |
| Uganda | 371 (61.1) |
| Reported Cough | 563 (92.8) |
| Reported Fever | 462 (76.1) |
| Failure to thrive | 122 (20.1) |
| Xpert Ultra positive result on sputum | 136 (22.4) |
| Culture on sputum positive for *M. tuberculosis* | 63 (10.4) |
| TB Classification |  |
| Confirmed TB | 147 (24.2) |
| Unconfirmed TB | 290 (47.8) |
| Unlikely TB | 170 (28.0) |
| Stool collection method (n=596) |  |
| Diaper | 159 (26.4) |
| Cup | 428 (71.1) |
| Other | 15 (2.5) |
| Stool processing method with valid result<sup>b</sup> |  |
| SPK (n=585) | 528 (90.3) |
| SOS (n=285) | 253 (88.8) |
| OSF (n=270) | 236 (87.4) |

a. N= 607 unless indicated
b. Introduction of SPK testing began first, followed by SOS and OSF, with respective totals noted

### Xpert Ultra Results

The three methods had a similar proportion of valid results, ranging from 87.4% for OSF to 90.3% for SPK (**Table 1**). As shown in **Table 2**, the proportion of *Mtb* positive results was similar across methods, ranging from 9.3% for OSF to 11.2% for SOS. The cycle threshold (CT) values among those with TB were lower in OSF (median 17.1, IQR 16.6-22.1) compared to SPK and SOS (median 20.6 and 19.8). Stool Xpert Ultra detected rifampin resistance in one child in South Africa using SPK, though this child was not tested with the other two methods.

**Table 2.** Summary of stool and sputum Xpert Ultra Results^1^.

|  | **SPK**<br>**(N=585)** | **SOS**<br>**(N=285)** | **OSF (N=270)<sup>2</sup>** | **Sputum Sample**<br>**(N=607)<sup>3,4</sup>** |
| --- | --- | --- | --- | --- |
| **MTB Positive, n (%)** | 55 (9.4) | 32 (11.2) | 25 (9.3) | 131 (21.6) |
| **Median CT value** | 20.6 (17.1-24.3) | 19.8 (16.7-25.4) | 17.1 (16.6-22.1) | - |
| **Trace** | 10 (18.2) | 7 (21.9) | 4 (16.7) | 36 (48) |
| **Very Low** | 14 (25.5) | 8 (25) | 3 (12.5) | 12 (16) |
| **Low** | 19 (34.6) | 11 (34.4) | 12 (50) | 7 (9.3) |
| **Medium** | 11 (20) | 5 (15.6) | 4 (16.7) | 11 (14.7) |
| **High** | 1 (1.8) | 1 (3.1) | 1 (4.2) | 9 (12) |
| **RIF resistant** | 1 (1.8) | 0 | 0 | 12 (16) |
| **MTB Negative, n (%)** | 473 (80.9) | 221 (77.5) | 211 (78.2) | 474 (78.1) |
| **Non-determinate results, n (%)** | 57 (9.7) | 32 (11.3) | 34 (12.6) | 2 (0.3) |
| **Invalid** | 41 (7.0) | 25 (8.8) | 13 (4.8) | 1 (0.2) |
| **Error** | 14 (2.4) | 7 (2.5) | 20 (7.4) | 1 (0.2) |
| **No Result** | 2 (0.3) | 0 | 1 (0.4) | 0 |

MTB: *M. tuberculosis*; RIF: Rifampin; SPK: Stool Processing Kit; SOS: Simple-One-Step; OSF: Optimized Sucrose Flotation
1. Summary of all testing, not limited to children who had all three stool processing methods, and is indicated by totals 2. One missing OSF semi-quantitative result 3. Median CT values not available for respiratory Xpert Ultra as results from South Africa came from a referral laboratory 4. Data on semi-quantitative level and rifampin resistance not available in South Africa, or if the child had sputum Xpert Ultra testing prior to enrollment (n=75)

**Table 2** also summarizes the Xpert Ultra results with sputum samples. Sputum samples were positive in 21.6% of children (131/607), with 64% (48/75) showing trace or very low semi-quantitative results. Only 2 children had non-determinate results.

### Accuracy of stool Xpert Ultra with centrifuge-free stool processing methods

The specificity of all three methods was high (97.1-98.2%), while sensitivity of the SPK, SOS and OSF methods was 36.9%, 38.6% and 31.3%, respectively, compared to the MRS (**Table 3**). Sensitivity was lower with the CRS (range 13.0-16.7%), similar compared to sputum Xpert Ultra (range 38.3-45%) and higher compared to sputum culture (range 55-77.3%). Across the methods, sensitivity was significantly higher when the sputum Xpert Ultra semi-quantitative level was Low or higher (95.8-100%), compared to Trace or Very Low (16.7-22.6%, p < 0.001 for all methods, **Supplemental Table 1**). In the intention-to-diagnose analysis, sensitivity ranged from 33.3-43.1% for SPK, 37.0-41.1% for SOS, and 28.1-38.0% for OSF. Specificity ranged from 88.7-98.4% for SPK, 84.0-97.6% for SOS and 83.9-97.5% for OSF (**Supplemental Table 2**).

**Table 3.**
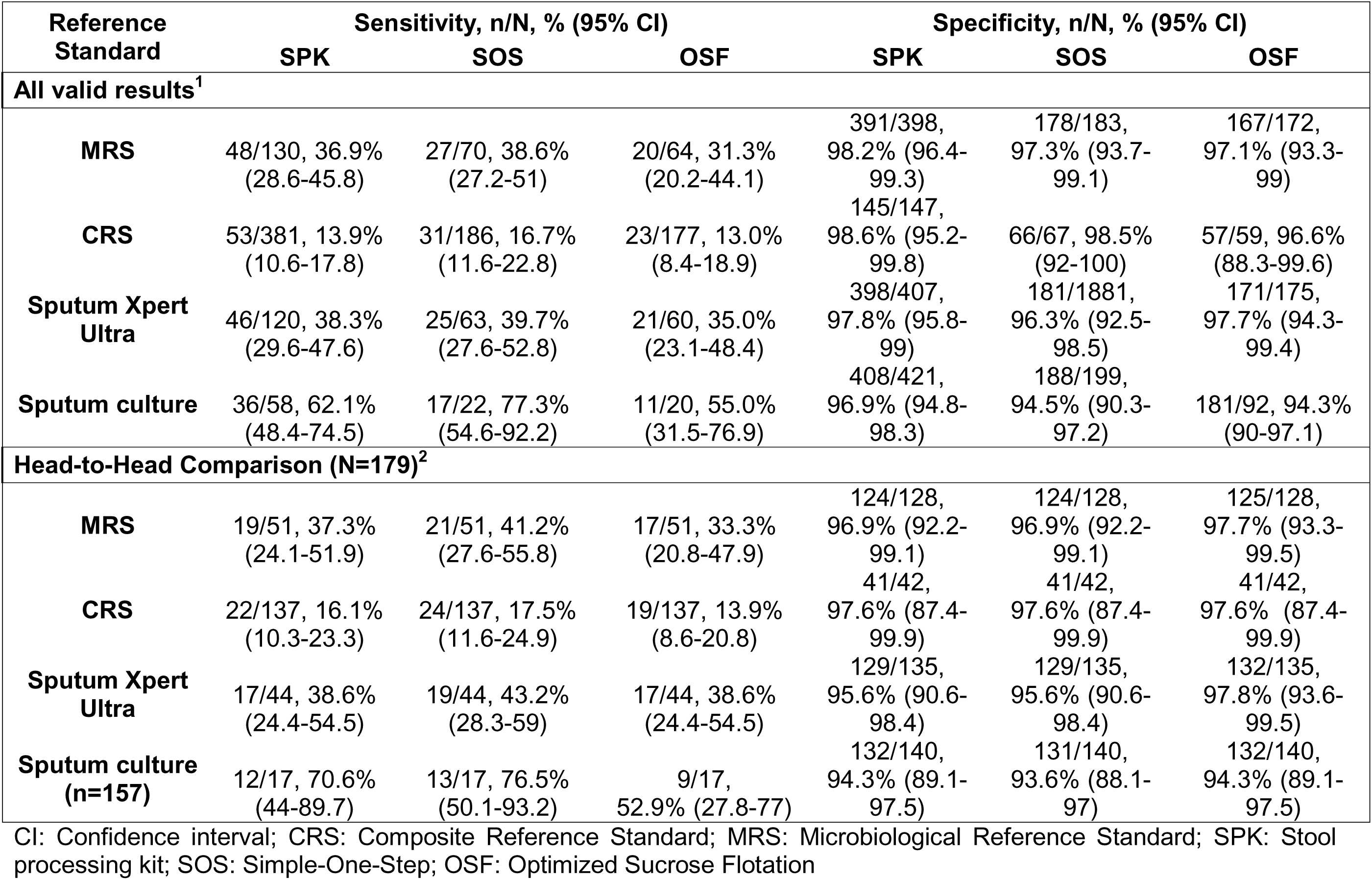
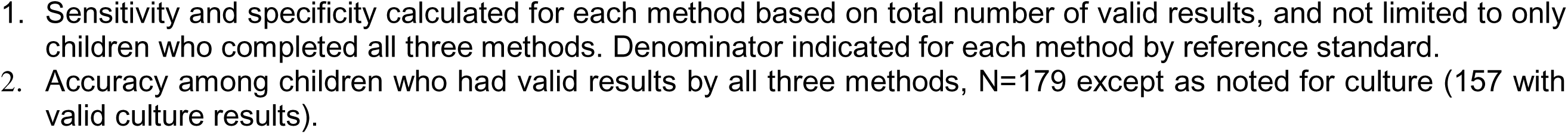
Diagnostic accuracy of stool Xpert Ultra, by centrifuge-free processing method.

| Reference Standard | Sensitivity, n/N, % (95% CI) |  |  | Specificity, n/N, % (95% CI) |  |  |
| --- | --- | --- | --- | --- | --- | --- |
|  | SPK | SOS | OSF | SPK | SOS | OSF |
| **All valid results<sup>1</sup>** |  |  |  |  |  |  |
| **MRS** | 48/130, 36.9%<br>(28.6-45.8) | 27/70, 38.6%<br>(27.2-51) | 20/64, 31.3%<br>(20.2-44.1) | 391/398,<br>98.2% (96.4-99.3) | 178/183,<br>97.3% (93.7-99.1) | 167/172,<br>97.1% (93.3-99) |
| **CRS** | 53/381, 13.9%<br>(10.6-17.8) | 31/186, 16.7%<br>(11.6-22.8) | 23/177, 13.0%<br>(8.4-18.9) | 145/147,<br>98.6% (95.2-99.8) | 66/67, 98.5%<br>(92-100) | 57/59, 96.6%<br>(88.3-99.6) |
| **Sputum Xpert Ultra** | 46/120, 38.3%<br>(29.6-47.6) | 25/63, 39.7%<br>(27.6-52.8) | 21/60, 35.0%<br>(23.1-48.4) | 398/407,<br>97.8% (95.8-99) | 181/1881,<br>96.3% (92.5-98.5) | 171/175,<br>97.7% (94.3-99.4) |
| **Sputum culture** | 36/58, 62.1%<br>(48.4-74.5) | 17/22, 77.3%<br>(54.6-92.2) | 11/20, 55.0%<br>(31.5-76.9) | 408/421,<br>96.9% (94.8-98.3) | 188/199,<br>94.5% (90.3-97.2) | 181/92, 94.3%<br>(90-97.1) |
| **Head-to-Head Comparison (N=179)<sup>2</sup>** |  |  |  |  |  |  |
| **MRS** | 19/51, 37.3%<br>(24.1-51.9) | 21/51, 41.2%<br>(27.6-55.8) | 17/51, 33.3%<br>(20.8-47.9) | 124/128,<br>96.9% (92.2-99.1) | 124/128,<br>96.9% (92.2-99.1) | 125/128,<br>97.7% (93.3-99.5) |
| **CRS** | 22/137, 16.1%<br>(10.3-23.3) | 24/137, 17.5%<br>(11.6-24.9) | 19/137, 13.9%<br>(8.6-20.8) | 41/42,<br>97.6% (87.4-99.9) | 41/42,<br>97.6% (87.4-99.9) | 41/42,<br>97.6% (87.4-99.9) |
| **Sputum Xpert Ultra** | 17/44, 38.6%<br>(24.4-54.5) | 19/44, 43.2%<br>(28.3-59) | 17/44, 38.6%<br>(24.4-54.5) | 129/135,<br>95.6% (90.6-98.4) | 129/135,<br>95.6% (90.6-98.4) | 132/135,<br>97.8% (93.6-99.5) |
| **Sputum culture (n=157)** | 12/17, 70.6%<br>(44-89.7) | 13/17, 76.5%<br>(50.1-93.2) | 9/17,<br>52.9% (27.8-77) | 132/140,<br>94.3% (89.1-97.5) | 131/140,<br>93.6% (88.1-97) | 132/140,<br>94.3% (89.1-97.5) |

CI: Confidence interval; CRS: Composite Reference Standard; MRS: Microbiological Reference Standard; SPK: Stool processing kit; SOS: Simple-One-Step; OSF: Optimized Sucrose Flotation
1. Sensitivity and specificity calculated for each method based on total number of valid results, and not limited to only children who completed all three methods. Denominator indicated for each method by reference standard. 2. Accuracy among children who had valid results by all three methods, N=179 except as noted for culture (157 with valid culture results).

Subgroup analysis is shown in **Supplemental Table 1.** Against the MRS, specificity was higher in Uganda for the SOS and OSF method. By age group, there was a trend towards lower sensitivity in children under 5 years old, and this was statistically significant for SOS (25.9% for children <5 years versus 66.7% for 10-14 years, p = 0.01). Sensitivity also tended to be higher in females, and was significantly higher for SPK (45.1% in females vs. 27.1% in males, p = 0.04).

### Head-to-head comparison of the stool processing methods

179 children had valid results from all three approaches (**Figure 1**). Comparing methods, sensitivity and specificity were similar regardless of the reference standard with overlapping 95% CIs (**Table 3**).

Against sputum culture, Xpert Ultra on sputum had higher sensitivity (82.4%, 95% CI 56.6-96.2) than stool Xpert Ultra in all three methods, but it was not statistically significant (**Table 4**). Specificity was significantly lower than stool Xpert Ultra across methods. When respiratory and stool Xpert Ultra testing were done concurrently, the sensitivity ranged from 82.4% (OSF) to 94.1% (SPK and SOS) against sputum culture. This represented an absolute increase of 17.6-23.5% compared to stool Xpert Ultra alone (**Table 4**), with a reduction in specificity of 11.4-12.1%. The addition of stool Xpert Ultra to sputum Xpert Ultra increased sensitivity by 0-11.8%, at a loss of specificity by 2.1-2.9%.

**Table 4.**
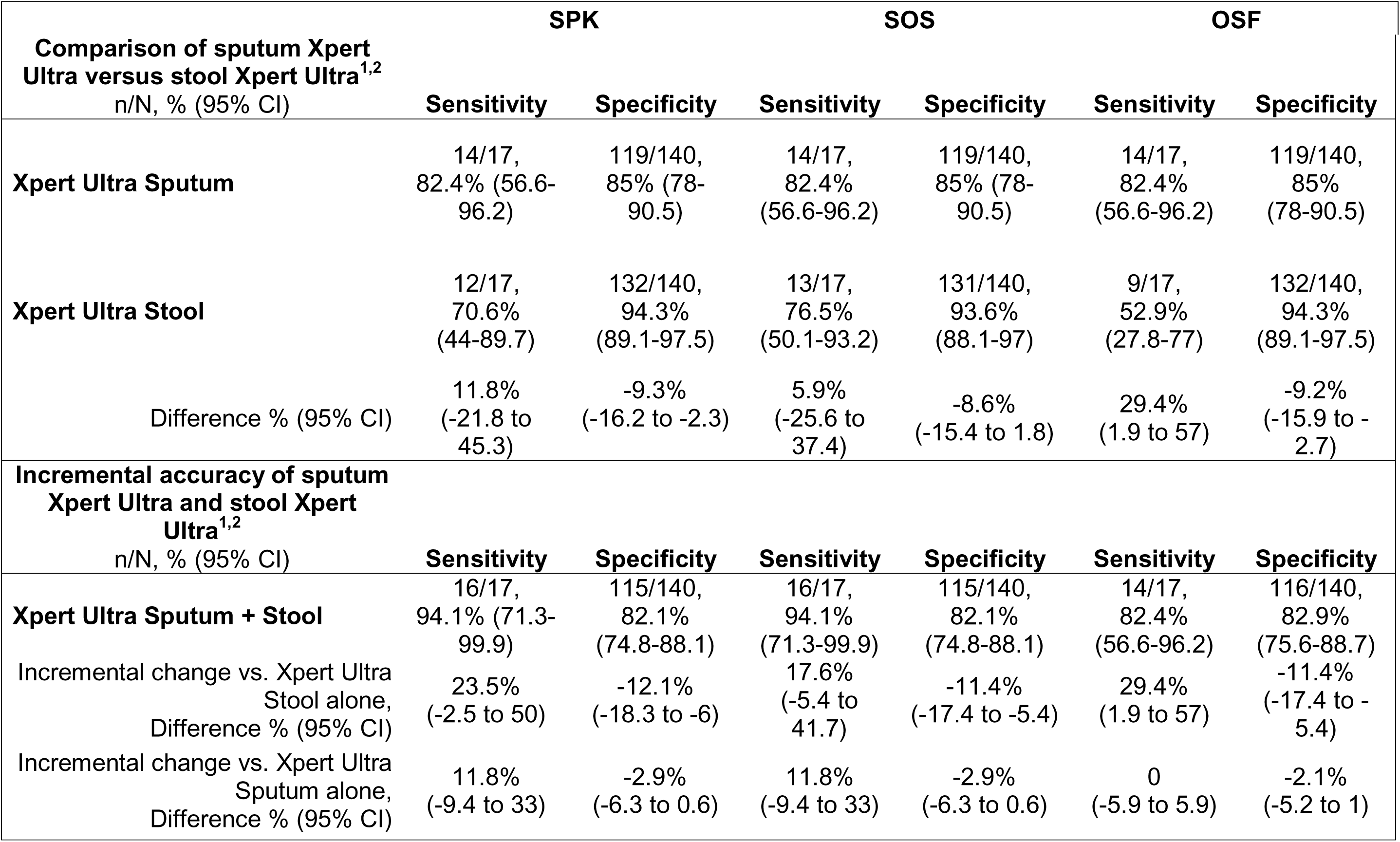
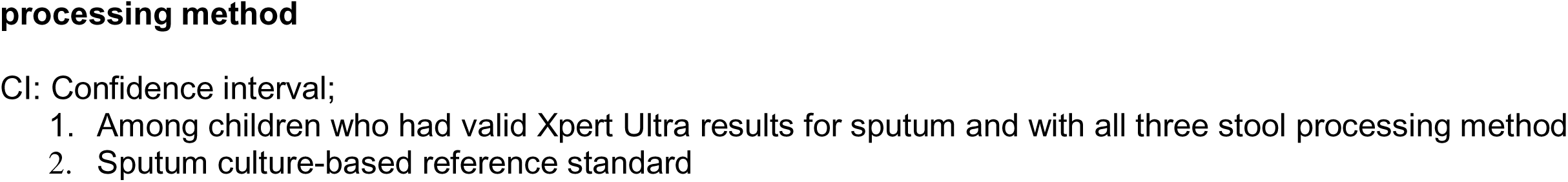
Comparison and incremental accuracy of Xpert Ultra with a sputum and stool specimen, by stool.

| Comparison of sputum Xpert Ultra versus stool Xpert Ultra<sup>1,2</sup><br>n/N, % (95% CI) | SPK |  | SOS |  | OSF |  |
| --- | --- | --- | --- | --- | --- | --- |
|  | Sensitivity | Specificity | Sensitivity | Specificity | Sensitivity | Specificity |
| **Xpert Ultra Sputum** | 14/17,<br>82.4% (56.6-96.2) | 119/140,<br>85% (78-90.5) | 14/17,<br>82.4% (56.6-96.2) | 119/140,<br>85% (78-90.5) | 14/17,<br>82.4% (56.6-96.2) | 119/140,<br>85% (78-90.5) |
| **Xpert Ultra Stool** | 12/17,<br>70.6% (44-89.7) | 132/140,<br>94.3% (89.1-97.5) | 13/17,<br>76.5% (50.1-93.2) | 131/140,<br>93.6% (88.1-97) | 9/17,<br>52.9% (27.8-77) | 132/140,<br>94.3% (89.1-97.5) |
| Difference % (95% CI) | 11.8%<br>(-21.8 to 45.3) | -9.3%<br>(-16.2 to -2.3) | 5.9%<br>(-25.6 to 37.4) | -8.6%<br>(-15.4 to 1.8) | 29.4%<br>(1.9 to 57) | -9.2%<br>(-15.9 to -2.7) |
| **Incremental accuracy of sputum Xpert Ultra and stool Xpert Ultra<sup>1,2</sup>**<br>n/N, % (95% CI) | **Sensitivity** | **Specificity** | **Sensitivity** | **Specificity** | **Sensitivity** | **Specificity** |
| **Xpert Ultra Sputum + Stool** | 16/17,<br>94.1% (71.3-99.9) | 115/140,<br>82.1% (74.8-88.1) | 16/17,<br>94.1% (71.3-99.9) | 115/140,<br>82.1% (74.8-88.1) | 14/17,<br>82.4% (56.6-96.2) | 116/140,<br>82.9% (75.6-88.7) |
| Incremental change vs. Xpert Ultra Stool alone,<br>Difference % (95% CI) | 23.5%<br>(-2.5 to 50) | -12.1%<br>(-18.3 to -6) | 17.6%<br>(-5.4 to 41.7) | -11.4%<br>(-17.4 to -5.4) | 29.4%<br>(1.9 to 57) | -11.4%<br>(-17.4 to -5.4) |
| Incremental change vs. Xpert Ultra Sputum alone,<br>Difference % (95% CI) | 11.8%<br>(-9.4 to 33) | -2.9%<br>(-6.3 to 0.6) | 11.8%<br>(-9.4 to 33) | -2.9%<br>(-6.3 to 0.6) | 0<br>(-5.9 to 5.9) | -2.1%<br>(-5.2 to 1) |

### Acceptability and usability of testing by laboratory staff

The survey was completed by 17 laboratory staff (5 from India, 7 from South Africa, and 5 from Uganda). The majority (12/17, 70.6%) had over three years of TB lab experience, but nearly half (8/17, 47.1%) had no prior experience with stool samples. All participants reported being comfortable handling stool samples and all agreed that stool testing with Xpert Ultra would be beneficial.

Usability is shown in **Figure 2**. For all three methods, the instructions were clear and posed minimal biosafety concern (>75% agreement). However, SOS was the least time-consuming, and most participants (75%) agreed it could be performed by non-laboratory staff at peripheral facilities with Xpert Ultra but without other laboratory infrastructure (94% agreement). Most respondents believed that all three methods could be performed at a peripheral health center if a microscopy laboratory was present.

**Figure 2.**
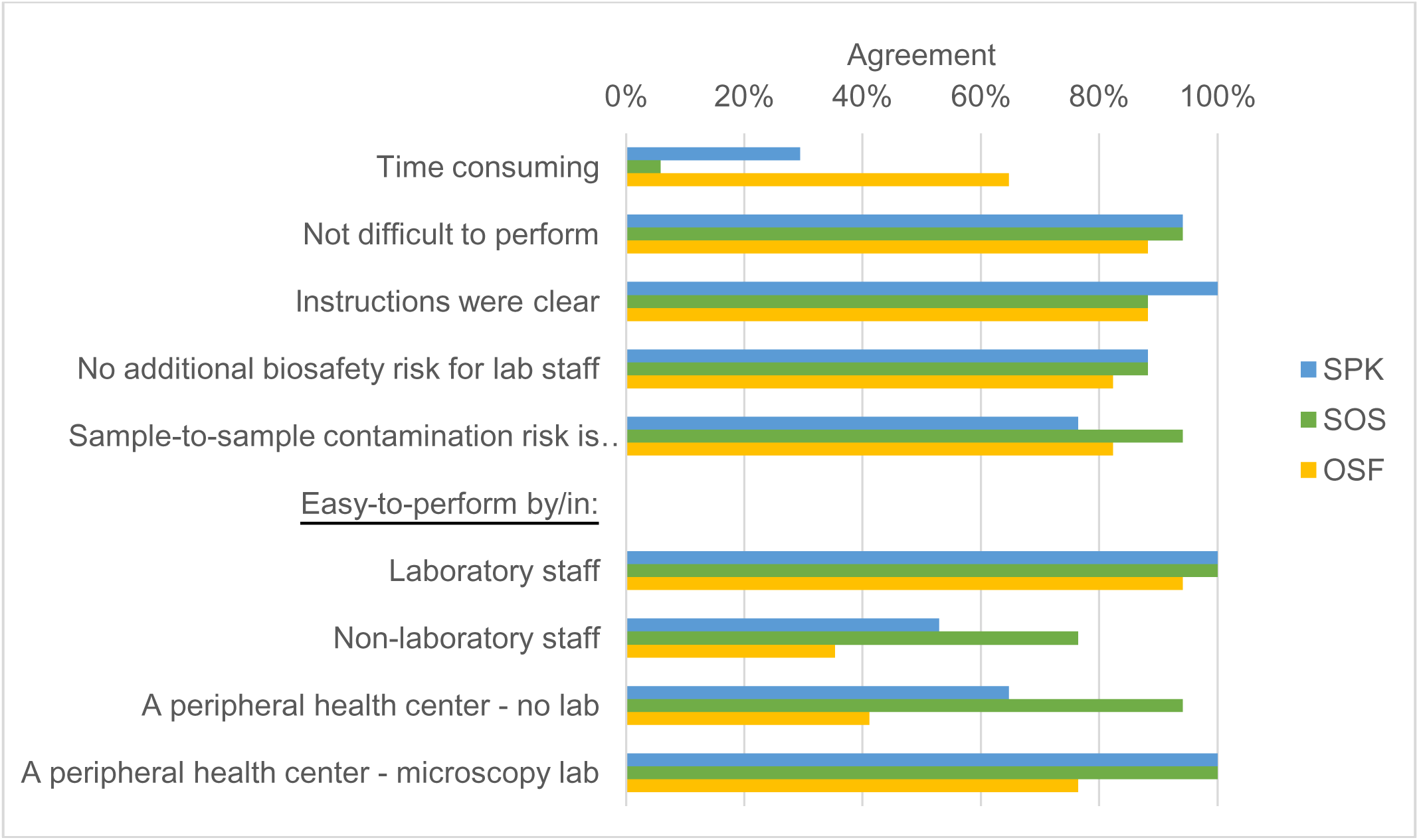
Usability of stool-based Xpert Ultra testing with centrifuge-free processing methods.

## DISCUSSION

Stool Xpert Ultra testing has the potential to increase access to molecular TB testing for children, especially in peripheral settings. In this multi-country, prospective evaluation of three stool processing methods for Xpert Ultra testing, we found that stool Xpert Ultra had high specificity, but detected only about a third of microbiologically confirmed cases with similar results across methods. Sensitivity doubled among children with culture-positive TB. Laboratory staff reported that all three methods were acceptable and usable, with SOS being the most practical for use by non-laboratory staff in peripheral facilities. However, ongoing gaps remain in improving TB diagnosis among children who are culture-negative and have paucibacillary disease.

Stool Xpert Ultra testing showed moderate sensitivity to detect culture-positive TB in children. This is consistent with studies that have shown similar stool and sputum Xpert Ultra accuracy in culture-positive TB [10, 14–16]. However, only 38-45% of children with positive sputum Xpert Ultra results were also positive by stool Xpert Ultra, which was lower than previous studies [10, 17–21]. One reason for this difference is that stool xpert performance has been shown to be heterogenous [14, 15], depending on collection and processing methods, age, setting, and co-morbidities. This is supported by the subgroup analysis where we noted differences by country, age group, and sex. Furthermore, both our study and prior studies found that bacterial burden was associated with stool *Mtb* detection [10, 17]. Our study had a higher proportion of trace or very low positive results on sputum Xpert Ultra compared to previous studies [5, 17], which could increase the likelihood of negative results on stool Xpert Ultra. Moreover, a companion stool Xpert Ultra study in Uganda and Zambia found higher sensitivity, but these children were mostly hospitalized and less likely to be TB contacts (28% versus 52% in our study), and so may have reflected later and more severe disease [22].

These findings suggest that while the accuracy of stool Xpert Ultra testing may be higher for culture-positive and more severe TB disease, its sensitivity is limited for children with paucibacillary disease [6]. This has important implications, because stool-based testing is particularly helpful in peripheral settings where children may present earlier, and in children under five who are more likely to have paucibacillary disease. We found that performing sputum Xpert Ultra in parallel with stool Xpert Ultra, or when stool Xpert Ultra is negative, increased absolute sensitivity by 17.6 to 29.4%. This is supported by other studies that have shown the combination of sputum and stool testing improved accuracy [17, 23]. Although specificity was lower for Xpert Ultra on sputum specimens using a culture-based reference standard, this likely reflects detection of culture-negative disease that has been previously reported in children [24, 25]. The addition of stool Xpert Ultra to sputum Xpert Ultra testing had a modest benefit. These findings informed a recent WHO Rapid Communication that recommended concurrent stool and respiratory molecular testing for childhood TB [26]. Further work is needed to assess how local TB prevalence and cost-effectiveness inform the benefit of combined testing.

All three centrifuge-free methods had a relatively large proportion of non-determinate results (9.7-12.6%), exceeding the WHO target product profile of <5% [27]. This was also observed during the development of these methods (7.8-10%) [9, 28, 29], and in past clinical studies [10, 17]. The reasons may be related to stool consistency, debris in the supernatant and presence of PCR inhibitors. Studies in Ethiopia and Vietnam using SOS found that validity increased as staff gained experience over time, though non-determinate results remained slightly higher than with sputum Xpert Ultra [10, 21]. In the intention-to-diagnose analyses, we found that including non-determinate results varied sensitivity by 3-7%, and specificity by 10-14%. Further improvements in processing and testing are needed to reduce the proportion of non-determinate results.

In the head-to-head comparison, the three methods performed similarly, and laboratory staff found them acceptable and usable. These findings contributed to the WHO guidelines endorsing stool for molecular testing in children [30]. SOS was the least time-consuming, and the most feasible for use in peripheral facilities without laboratory infrastructure. Although SPK and OSF were at earlier stages of development, the findings highlight the preference for methods that require fewer supplies and processing steps. SPK development has not continued due to the similar performance and additional supplies required. SOS is being implemented in high TB burden settings, and data suggests increased access to testing and pediatric TB notifications [31], further highlighting the importance of non-invasive sample types to increase access to TB testing for children.

This study is the largest, prospective evaluation comparing stool processing methods for children in multiple high TB-burden settings. We utilized a standardized protocol and reference standards across sites, and our randomization procedure for stool testing minimized bias in the head-to-head comparison. However, we also acknowledge several limitations. SPK had a larger sample size due to earlier introduction; its diagnostic accuracy estimates may be more reliable compared to SOS and OSF, though the comparative analysis included only participants who underwent all three tests. We had few drug-resistant cases and further assessment is needed. Additionally, we did not include caregivers in our acceptability and usability survey, but a recent study found that most caregivers preferred stool over respiratory testing [10].

## CONCLUSIONS

Three stool processing methods achieved similar accuracy with Xpert Ultra, and performed best among children who are *Mtb* culture-positive. All methods were acceptable, but SOS was the most feasible to be implemented in peripheral facilities without a laboratory. Centrifuge-free stool processing methods may increase access to Xpert Ultra for children in those settings, but sensitivity is lower than sputum Xpert Ultra in children with culture-negative disease. Additional implementation and economic evaluations are needed to assess the benefits of stool testing when sputum Xpert Ultra testing in children is feasible. Stool testing alone may not eliminate the diagnostic gap for pediatric TB, but could play an important role as part of a comprehensive package of TB diagnostic approaches and highlights the need to develop more sensitive assays on easily accessible samples to detect TB in children.

## Data Availability

De-identified data used for the analysis is available on request.

## ACKNOWLEDGEMENTS

We thank the study participants, their caregivers and clinical and research staff at each site. We also thank FIND staff Flavio Ambrogiani, Aradhana Chauhan, Loghanathan Prabakaran, Sunita Singh, and Rita Szekely for their support. We greatly appreciate Cepheid for donating Xpert Ultra cartridges and loaning GeneXpert machines. De-identified data used for the analysis is available on request.

For author contributions: conceptualization was by DJ, PN, MPN, HJZ, RL, AB, AC, MR, EW, CMD, methodology by DJ, PN, MPN, MB, ML, PD, DA, PB, and CMD, investigation by DJ, MPN, RC, PW, HJZ, LW, RL, UBS, AB, AT, AM, AC, and EW, analysis by DJ, RC, and LW, writing original draft by DJ, review and edits by all authors, and resources by DJ, AC, MR, EW, and CMD.

The work was supported by the National Institutes of Health, through the National Institute of Allergy and Infectious Diseases [U01AI152087 to AC and CMD; and R01AI131617 to DA], and the National Heart, Lung, and Blood Institute [K23HL153581 to DJ]. The content is solely the responsibility of the authors and does not necessarily represent the official views of the NIH. FIND provided funding to all the sites and the SPK supplies. Cepheid provided in-kind support of Xpert Ultra cartridges and GeneXpert machines, but had no role in the study design, analysis, interpretation and decision to publish.

For potential conflicts of interest, MPN, DA, PB and FIND were involved in the development of SPK, PD and ET for SOS, and MB and ML for OSF. DA holds patents related to tuberculosis detection and drug treatment, and receives royalty payments for one or more of these patents that have been licensed by Rutgers University to Cepheid. The other authors declare no conflicts of interest.

