## Supplemental Figure 1 for "Stool processing methods for Xpert Ultra testing in childhood tuberculosis: A prospective, multi-country accuracy study"

### Supplemental Figure 1. Overview of Stool Processing Methods for Xpert Ultra testing

#### A. Stool Processing Kit (SPK)

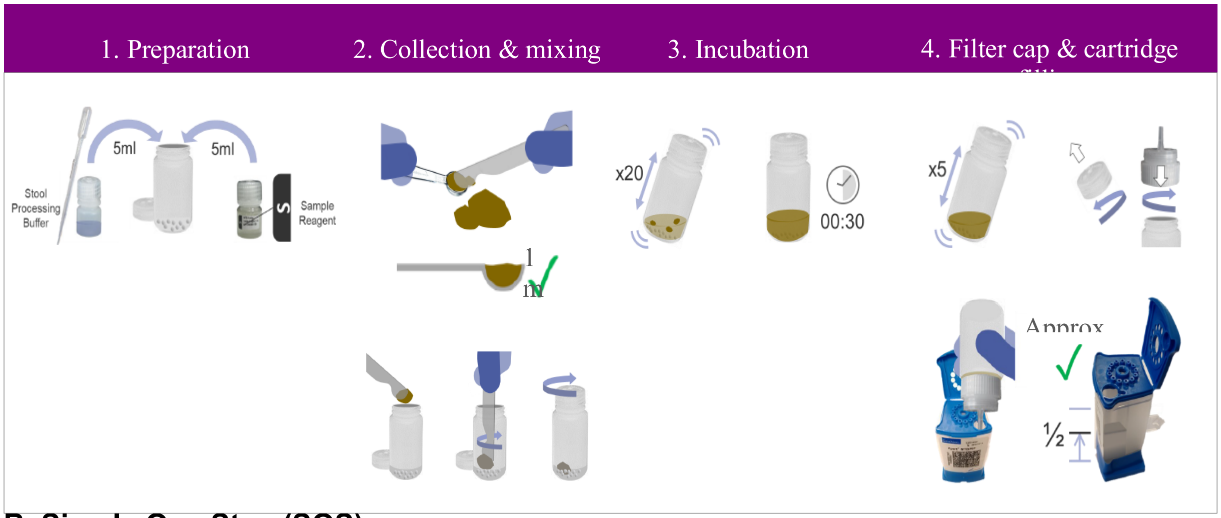


#### B. Simple One Step (SOS)

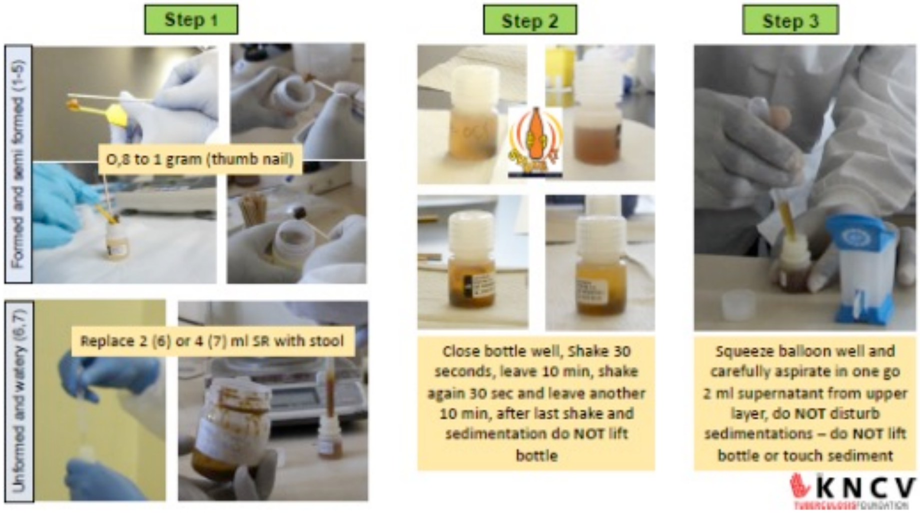


#### C. Optimized Sucrose Flotation (OSF)

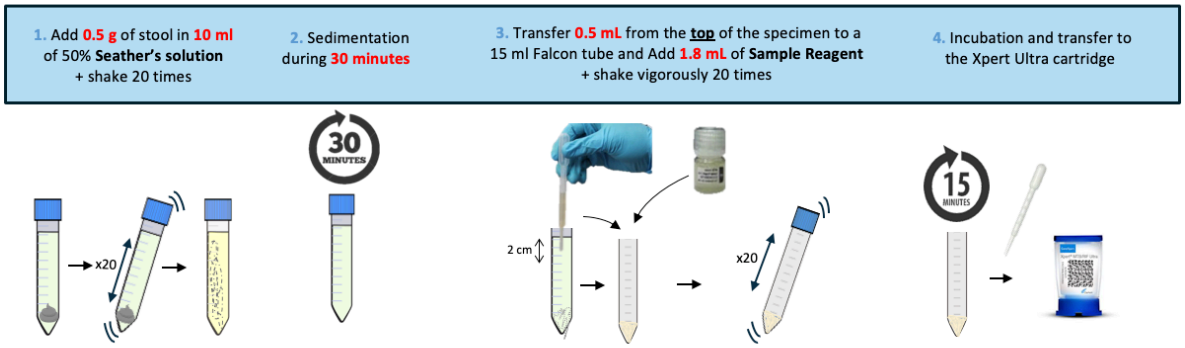
