## Supplemental Table 1 for "Stool processing methods for Xpert Ultra testing in childhood tuberculosis: A prospective, multi-country accuracy study"

**Supplemental Table 1. Diagnostic accuracy of each centrifuge-free stool processing method by subgroup**

| | Sensitivity, n/N, % (95% CI)[1,4] | | | | | | Specificity, n/N, % (95% CI)[1,4] | | | | | |
|---|---|---|---|---|---|---|---|---|---|---|---|---|
| | SPK | p-value[2] | SOS | p-value | OSF | p-value | SPK | p-value | SOS | p-value | OSF | p-value |
| **Country** | | | | | | | | | | | | |
| **Uganda** | 25/56, 44.6% (31.3-58.5) | 0.28 | 12/30, 40% (22.7-59.4) | 1 | 8/26, 30.8% (14.3-51.8) | 1 | 281/284, 98.9% (96.9-99.8) | 0.19 | 152/153, 99.4% (96.4-100) | 0.01 | 138/139, 99.3% (96.1-100) | 0.01 |
| **South Africa** | 14/45, 31.1% (18.2-25.3) | | 2/6, 33.3% (4.3-77.7) | | 1/5, 20% (0.5 -71.6) | | 109/113, 96.5% (91.2-99) | | 27/30, 90% (73.5-97.9) | | 27/30, 90% (73.5-97.9) | |
| **India** | 9/29, 31.0% (15.3-50.8) | | 13/34, 38.2% (22.2-56.4) | | 11/33, 33.3% (18-51.8) | | 17/17, 100% (80.5-100) | | 14/15, 93.3% (68.1-99.8) | | 16/17, 94.1% (71.3-99.9) | |
| **Age Group** | | | | | | | | | | | | |
| **<5 years** | 19/60, 31.7% (20.3-45) | 0.12 | 7/27, 25.9% (11.1-46.3) | 0.01 | 5/25, 20% (6.8-40.7) | 0.17 | 252/259, 97.3% (94.5-98.9) | 0.14 | 120/124, 96.8% (91.9-99.1) | 0.29 | 118/121, 97.5% (92.9-99.5) | 0.61 |
| **5-9 years** | 14/42, 33.3% (19.6-49.5) | | 6/22, 27.3% (10.7-50.2) | | 6/20, 30% (11.9-54.3) | | 113/113, 100% (96.8-100) | | 53/53, 100% (93.3-100) | | 45/46, 97.8% (88.5-99.9) | |
| **10-14 years** | 15/28, 53.6% | | 14/21, 66.7% | | 9/19, 47.4% | | 42/42, 100% | | 20/21, 95.2% | | 18/19, 94.7% | |

| | | | | | | | | | | | | |
|---|---|---|---|---|---|---|---|---|---|---|---|---|
| | (33.9-72.5) | | (43-85.4) | | (24.4-71.1) | | (91.6-100) | | (76.2-99.9) | | (74-99.9) | |
| **Sex** | | | | | | | | | | | | |
| **Female** | 32/71, 45.1% (33.2-57.3) | 0.04 | 18/44, 40.9% (26.3-56.8) | 0.6 | 14/42, 33.3% (19.6-49.5) | 0.62 | 189/190, 99.5% (97.1-100) | 0.13 | 86/88, 97.7% (92-99.7) | 1 | 74/76, 97.4% (90.8-99.7) | 1 |
| **Male** | 16/59, 27.1% (16.4-40.3) | | 9/26, 34.6% (17.2-55.7) | | 6/22, 27.3% (10.7-50.2) | | 218/224, 97.3% (94.3-99) | | 107/110, 97.3% (92.2-99.4) | | 107/110, 97.3% (92.2-99.4) | |
| **HIV status** | | | | | | | | | | | | |
| **HIV Negative** | 35/107, 32.7% (24-42.5) | 0.13 | 15/52, 28.9% (17.1-43.1) | 0.63 | 12/45, 26.7% (14.6-41.9) | 1 | 334/340, 98.2% (96.2-99.3) | 1 | 151/156, 96.8% (92.7-99) | 1 | 136/140, 97.1% (92.8-99.2) | 1 |
| **HIV Positive** | 7/13, 53.9% (25.1-80.8) | | 2/5, 40% (86.3-100) | | 1/5, 20% (0.51-71.6) | | 60/61, 98.4% (91.2-100) | | 25/25, 100% (86.3-100) | | 25/28, 96.4% (81.7-99.9) | |
| **Nutritional Status** | | | | | | | | | | | | |
| **Not Underweight** | 15/51, 29.4% (17.5-43.8) | 0.15 | 7/24, 29.2% (12.6-51.1) | 0.31 | 5/22, 22.7% (7.8-45.4) | 0.29 | 211/214, 98.6% (96-99.7) | 0.72 | 94/97, 96.9% (91.2-99.4) | 0.68 | 95/97, 97.9% (92.7-99.7) | 0.67 |
| **Underweight** | 33/79, 41.8% (20.8-53.4) | | 20/46, 43.5% (28.9-58.9) | | 15/42, 35.7% (21.6-52) | | 196/200, 98% (95-99.5) | | 99/101, 98% (93-99.8) | | 86/89, 96.6% (90.5-99.3) | |
| **Respiratory Xpert Ultra Semi-quantitative Level[3]** | | | | | | | | | | | | |

| | | | | | | |
|---|---|---|---|---|---|---|
| **Trace or Very low** | 7/37, 18.9% (8-35.2) | <0.001 | 7/31, 22.6% (9.6-41.1) | <0.001 | 5/30, 16.7% (5.6-34.7) | <0.001 |
| **Low or Higher grade** | 23/24, 95.8% (78.9-99.9) | | 15/15, 100% (78.2-100) | | 14/14, 100% (76.8-100) | |

1. According to the microbiological reference standard
2. Chi-squared or Fisher exact testing
3. Among those with Confirmed TB, limited to sensitivity comparison
4. Sensitivity and specificity calculated for each method based on total number of valid results, and not limited to only children who completed all three methods. Denominator indicated for each method by reference standard
