## Supplemental Table 2 for "Stool processing methods for Xpert Ultra testing in childhood tuberculosis: A prospective, multi-country accuracy study"

**Supplemental Table 2. Intention-to-diagnose analysis to assess the accuracy of three centrifuge-free stool processing methods, according to a microbiological reference standard**

| | Sensitivity, n/N, % (95% CI) | | | Specificity, n/N, % (95% CI) | | |
|---|---|---|---|---|---|---|
| | **SPK** | **SOS** | **OSF** | **SPK** | **SOS** | **OSF** |
| **Valid results only** | 48/130, 36.9% (28.6-45.8) | 27/70, 38.6% (27.2-51) | 20/64, 31.3% (20.2-44.1) | 391/398, 98.2% (96.4-99.3) | 178/183, 97.3% (93.7-99.1) | 167/172, 97.1% (93.3-99) |
| **Non-determinate[1] results as positive** | 62/144, 43.1% (34.8-51.6) | 30/73, 41.1% (29.7-53.2) | 27/71, 38.0% (26.8-50.3) | 391/441, 88.7% (85.3-91.5) | 178/212, 84.0% (78.3-88.6) | 167/199, 83.9% (78.1-88.7) |
| **Non-determinate results as negative** | 48/144, 33.3% (25.7-41.7) | 27/73, 37.0% (26-49.1) | 20/71, 28.1% (18.1-40.1) | 434/441, 98.4% (96.8-99.4) | 207/212, 97.6% (94.6-99.2) | 194/199, 97.5% (94.2-99.2) |

CI: Confidence Interval; SPK: Stool processing kit; SOS: Simple-One-Step; OSF: Optimized Sucrose Flotation

1. Non-determinate defined as invalid, error or no-result.
